# Concurrent Gabapentin and Dihydropyridine Calcium Channel Blocker Use and Risk of Incident Dementia: An Active-Comparator Cohort Study

**DOI:** 10.64898/2026.02.06.26345763

**Authors:** James W. Green, Luciana Mascarenhas Fonseca, Michal Schnaider Beeri, Sharon Sanz Simon, James Scott Parrott, Branimir Ljubic, Michael Schulewski, Barbara Tafuto, Suril Gohel

## Abstract

**Importance:** Gabapentin and dihydropyridine calcium channel blockers (DHP CCBs) are commonly prescribed together in older adults: gabapentin reaches more than 15.5 million U.S. patients annually, and DHP CCBs such as amlodipine are a first-line antihypertensive class. Both classes act on neuronal calcium signaling, raising the question of whether their combined use is associated with greater cognitive risk than gabapentin alone.

**Objective:** To examine whether concurrent CCB use, and DHP CCB use in particular, modifies the association between gabapentin initiation and incident dementia in adults with hypertension.

**Design, Setting, and Participants:** Active-comparator new-user cohort study within the Rutgers Clinical Research Data Warehouse (2018–2024), encompassing 405 clinical sites in northern New Jersey. Participants were adults aged 40 years or older with hypertension who initiated gabapentin or pregabalin. Data were analyzed from January through December 2025.

**Exposures:** Gabapentin vs pregabalin initiation, with pre-specified stratification by baseline CCB exposure and CCB subtype (DHP vs non-DHP).

**Main Outcomes and Measures:** Incident dementia (ICD-10 F00–F03, G30). The pre-specified primary analysis used inverse probability of treatment weighting with time-varying CCB exposure; secondary analyses examined CCB subtype specificity, dementia subtypes, falsification outcomes, and competing risk of death.

**Results:** Among 33,791 patients (mean [SD] age, 69.9 [11.5] years; 57.1% female; 502 dementia events; median follow-up, 1.22 years), gabapentin initiation was associated with higher dementia incidence among patients also taking a CCB than among those not taking a CCB (time-varying HR, 1.52 [95% CI, 1.23–1.89]; baseline-stratified HR, 2.22 [95% CI, 1.42–3.47]). The signal was directionally larger with DHP CCBs. Median time to dementia diagnosis was 240 days; subtype estimates were hypothesis-generating.

**Conclusions and Relevance:** In this active-comparator cohort of older adults with hypertension, concurrent CCB use — particularly with DHP agents — was associated with substantially higher gabapentin-related dementia incidence than gabapentin without CCB exposure. The short event latency suggests the signal may partly reflect drug-induced cognitive impairment, which may improve with deprescribing — a clinically actionable possibility. CCB-subtype and dementiasubtype estimates are hypothesis-generating; confirmatory replication in Medicare fee-for-service claims and the NIH All of Us Research Program is underway.

**Key Points:** *Question:* In older adults with hypertension, is concurrent calcium channel blocker (CCB) use associated with higher gabapentin-related dementia incidence than gabapentin alone?

*Findings:* In an active-comparator cohort of 33,791 adults, gabapentin initiation (vs pregabalin) was associated with higher dementia incidence among patients also taking a CCB than among those not taking a CCB (time-varying HR, 1.52 [95% CI, 1.23–1.89; *P* = .0003]; baselinestratified HR, 2.22 [95% CI, 1.42–3.47]; interaction *P* = .004). The signal was directionally larger among DHP CCB users (HR, 3.20) and concentrated in Alzheimer disease (HR, 4.53) and unspecified dementia (HR, 2.29), with a null finding for vascular dementia (HR, 0.81).

*Meaning:* Concurrent gabapentin and DHP CCB use identifies a potentially modifiable cognitive safety signal in older adults; the short event latency raises the possibility that part of the affected cognition may be at least partially reversible after deprescribing. CCB-subtype and dementiasubtype findings are hypothesis-generating and require replication in larger cohorts; independent replication efforts in broader real-world populations are ongoing.

## Introduction

Gabapentin and calcium channel blockers (CCBs) are two of the most widely prescribed medication classes in adults aged 65 years and older. Gabapentin prescriptions in the United States grew from 24 million in 2010 to nearly 59 million by 2024, reaching 15.5 million patients annually, with the highest prescribing rates in older adults.^1^·^2^ Originally approved for epilepsy and postherpetic neuralgia, its use has expanded to diabetic neuropathy, fibromyalgia, and a range of off-label indications.^3^· Dihydropyridine CCBs such as amlodipine, by contrast, are a first-line antihypertensive class. Because adults with chronic pain conditions also have high rates of comorbid hypertension (pooled OR, 1.66; 95% CI, 1.28–2.15),^23^ co-prescription of gabapentin with a CCB is common in everyday geriatric practice. Whether this very common combination carries cognitive risk beyond gabapentin alone has not been examined.

Recent observational studies have linked gabapentin use to higher dementia incidence. Patients with chronic low back pain receiving 6 or more gabapentin prescriptions had 29% higher dementia risk than nonusers (RR, 1.29; 95% CI, 1.05–1.57), and gabapentin initiation has been associated with neurocognitive and behavioral changes in older adults with existing cognitive impairment. However, these studies compared gabapentin users with nonusers — a design susceptible to confounding by indication — and none examined whether the very common coprescription of gabapentin with other CNS-active or calcium-modulating medications modifies the association.

There is biological reason to ask this specific question. Gabapentin binds the α δ subunit of voltage-gated calcium channels and reduces presynaptic calcium influx. · ·^1^ ·^11^ DHP CCBs penetrate the central nervous system and block postsynaptic L-type calcium channels critical for hippocampal long-term potentiation,^12^·^13^·^1^ ·^1^ ·^1^ ·^1^ ·^1^ ·^1^ a premise directly supported by recent in vivo evidence that amlodipine crosses the blood-brain barrier at pharmacologically relevant concentrations.^1^ The convergence of presynaptic α δ blockade and postsynaptic Ltype blockade may impair calcium-dependent synaptic plasticity beyond what either drug produces in isolation. By contrast, non-DHP CCBs such as verapamil show greater cardiac selectivity and substantially less CNS penetration,^12^·^1^ and the DREAM consortium independently reported higher dementia risk among DHP CCB users than among hydrochlorothiazide users.^2^ These observations motivate a specific empirical question rather than a class-wide one: does CCB co-medication, and DHP CCB co-medication in particular, modify the gabapentin–dementia association?

We addressed this question using an active-comparator new-user design^2^ ·^2^ ·^2^ ·^2^ structured as a target trial emulation^2^ within a multi-site EHR cohort of adults with hypertension. Pregabalin served as the active comparator: it shares gabapentin’s α δ binding target and clinical indications but has linear pharmacokinetics with ≥90% bioavailability, in contrast to gabapentin’s saturable absorption and 20–30% interpatient variability.^22^ Pre-specified analyses examined CCB exposure as an effect modifier, DHP versus non-DHP subtype specificity, and dementia subtypes; methodological details are reported in the Methods. If confirmed, the signal examined here would identify a very common and potentially modifiable medication-related contributor to cognitive decline in older adults — addressable through prescriber awareness and medication review rather than through new therapeutics.

## Methods

### Study Design and Data Source

We used an active-comparator new-user design^2^ ·^2^ ·^2^ ·^2^ structured as a target trial emulation^2^ ·^31^ within the Rutgers Clinical Research Data Warehouse (CRDW; 2018– 2024).^2^ ·^3^ Table 1 maps the target-trial components to their observational analogs. The CRDW encompasses 405 clinical facilities within the Robert Wood Johnson Health System in northern New Jersey, containing approximately 4 million patient records. This study used completely de-identified data and was determined to be Non-Human Subject Research by the Rutgers University Institutional Review Board (Non-Human Research Self-Certification; PI: Branimir Ljubic; Project: Hypertension — Data Analytics and Computer Science Modeling; certified August 15, 2024). Informed consent was not required. We followed STROBE guidelines for reporting.^32^

**Table 1.** Emulated Target Trial Protocol.

| **Protocol Component** | **Target Trial** | **CRDW Emulation** |
| --- | --- | --- |
| **Eligibility criteria** | Adults ≥40 years with hypertension, no prior dementia, initiating a new gabapentinoid prescription | Same: ICD-10 I10–I16, age ≥40 years, 180-day baseline period, no prior dementia in EHR |
| **Treatment strategies** | 1. Initiate gabapentin; 2. Initiate pregabalin (active comparator with shared mechanism and indications) | Same: first gabapentinoid prescription = index date (newuser design) |
| **Treatment assignment** | Random allocation to gabapentin or pregabalin | Non-random; emulated via IPTW conditional on 34 baseline covariates; randomization assumed conditional on measured confounders |
| **Pre-specified effect modifier** | Baseline DHP CCB co-prescription (subtype specificity test: DHP vs non-DHP) | Any CCB prescription in 180-day pre-index period; DHP (amlodipine, nifedipine) vs non-DHP (verapamil, diltiazem) |
| **Causal contrast** | Intention-to-treat (ITT) effect of gabapentinoid initiation on incident dementia; pre-specified subgroup interaction by CCB status | Observational analog of ITT effect; time-varying CCB analysis as primary estimand (prespecified); baseline-stratified as sensitivity |
| **Follow-up** | From treatment assignment to incident dementia, death, or administrative censoring | From index date to incident dementia (ICD-10 F00–F03, G30), last encounter, or December 31, 2024 |
| **Statistical analysis** | Weighted survival analysis with pre-specified subgroup interaction; robust standard errors | IPTW Cox with robust sandwich SE; multiplicative interaction term; time-varying CCB as primary estimand per prespecified protocol |

Adults aged 40 years or older with diagnosed hypertension (ICD-10 I10–I16) who initiated gabapentin or pregabalin were eligible. The index date was the first gabapentinoid prescription. We required a 180-day baseline period with at least one healthcare encounter. Patients with any prior dementia diagnosis recorded in the EHR before the index date were excluded to ensure incident outcomes; a sensitivity analysis extending this window to 365 days yielded zero additional exclusions.

### Exposures

The primary exposure was gabapentin versus pregabalin initiation. Pregabalin shares gabapentin’s mechanism of action (α δ subunit binding), therapeutic indications, and prescriber population.^33^·^22^·^2^ Baseline CCB exposure was defined as any CCB prescription in the 180-day pre-index period, further classified as DHP (amlodipine, nifedipine; other DHP agents such as felodipine were captured in the any-CCB stratum but not in subtype-specific analyses, accounting for 61 of 6,146 CCB-exposed patients) or non-DHP (verapamil, diltiazem).

### Outcomes

The primary outcome was incident dementia, defined using a standard validated pharmacoepidemiologic ICD-10 code set (F00–F03, G30) consistent with prior claims-based dementia research.^3^ ·^3^ ·^3^ ·^3^ Secondary outcomes included dementia subtypes: F03 (unspecified dementia), G30 (Alzheimer disease), and F01 (vascular dementia). Two prespecified outcome sensitivity analyses addressed ICD-10 code reliability. First, a two-code definition required two dementia ICD codes recorded on separate dates within 60 days of each other, with the first code date assigned as the outcome date; this approach is associated with a positive predictive value exceeding 95% in validation studies.^3^ ·^3^ ·^3^ ·^3^ Second, an Rxconfirmed definition required an ICD dementia code plus a prescription for a cholinesterase inhibitor or memantine (donepezil, galantamine, rivastigmine, memantine) within 180 days, with the ICD code date as the outcome date; this definition reflects clinician commitment to the diagnosis sufficient to initiate pharmacotherapy.

### Covariates

Baseline covariates included demographics (age, sex [as recorded in the EHR], race), comorbidities (diabetes, cardiovascular disease, stroke, chronic kidney disease, depression, anxiety, atrial fibrillation, heart failure, insomnia, obesity, cancer, chronic pain, neuropathy, epilepsy), healthcare utilization (encounters, emergency visits, hospitalizations), and concomitant medications (antihypertensives, statins, opioids, benzodiazepines, SSRIs, SNRIs, proton pump inhibitors). Anticholinergic burden was quantified using the Anticholinergic Cognitive Burden (ACB) scale (2012 update; Aging Brain Care) applied to all baseline medications and included as a continuous covariate in the propensity score model at the time-fixed baseline, given its independent association with dementia risk.

### Statistical Analysis

Propensity scores for gabapentin initiation were estimated by logistic regression on 34 baseline covariates including anticholinergic burden (ACB). Stabilized inverse probability of treatment weights targeting the average treatment effect in the treated (ATT) were truncated at the 1st and 99th percentiles.^2^ ·^2^ ·^2^ ·^3^ ·^3^ · · ^1^· ^2^ Covariate balance was assessed using standardized mean differences (SMD); all 34 covariates achieved balance after IPTW. Cox proportional hazards models with robust sandwich standard errors estimated hazard ratios with multiplicative interaction terms for CCB exposure status. The proportional hazards assumption was evaluated using a time-interaction test on the gabapentin × log(t) term. The time-varying CCB analysis was pre-specified as the primary estimate; the baseline-stratified analysis is reported as a sensitivity analysis. Competing risk of death was addressed via Fine-Gray sub distribution hazard models combining IPTW weights with Fine-Gray redistribution weights and robust standard errors. No analysis plan was registered prior to data lock; the primary interaction analysis, dementia subtype analyses, lag analyses, falsification outcomes, duloxetine and non-α δ comparator analyses, and intrasample replication were prespecified before analysis, while the formal DHP-versus-nonDHP contrast, proportional hazards assessment, effective sample size analysis, and Fine-Gray competing risk analysis were added during peer review and are labeled as such in the Results.

### Validation Framework

Validation analyses included pre-index symptom balance, lag analyses, falsification outcomes, comparator analyses (non-α δ anticonvulsants and duloxetine), comorbidity stratification, and intrasample replication. Additional analyses are reported in the Supplement (eTables 1–5, eFigure 1).

**Figure 1.** Study Design and Cohort Composition. Integrated study design panel combining STROBE-style flow diagram (left) with cohort composition summary (right). Source population: adults aged 40 or older with hypertension and gabapentinoid prescription, Rutgers CRDW, 2018–2024. Final analytic cohort: N = 33,791 (502 dementia events). Gabapentin initiators n = 28,058 (83.0%); pregabalin initiators n = 5,733 (17.0%). CCB exposure at baseline: 18.2% (n = 6,146), predominantly DHP (84.6%). Primary outcome: CCB users HR = 2.22 (P = .0005; interaction P = .004); non-CCB users HR = 1.15 (P = .072).

## Results

### Study Population

The final analytic cohort included 33,791 patients: 28,058 gabapentin initiators (83.0%) and 5,733 pregabalin initiators (17.0%) (Figure 1). Mean age was 69.9 years (SD, 11.5), 57.1% were female, and the population was ethnoracially diverse (47.6% White, 19.0% Black, 26.3% Other race, 4.0% Asian). Baseline CCB exposure was present in 6,146 patients (18.2%), predominantly DHP CCBs (84.6% of CCB users). After IPTW, all 34 covariates achieved adequate balance (SMD < 0.10), including SNRI use (SMD, 0.067), anticholinergic burden (SMD, 0.025), atrial fibrillation (SMD, 0.009), and heart failure (SMD, 0.021); residual channeling by indication cannot be fully excluded (Table 1).

**Table 1.** Characteristics of Gabapentin and Pregabalin Initiators.

| Characteristic | Gabapentin (n = 28,058) | Pregabalin (n = 5,733) | Adjusted SMD |
| --- | --- | --- | --- |
| Age, mean (SD), y | 70.0 (11.6) | 69.6 (11.2) | 0.000 |
| Female sex, n (%) | 15,965 (56.9) | 3,331 (58.1) | 0.004 |
| White race, n (%) | 13,215 (47.1) | 2,889 (50.4) | 0.075 |
| Chronic kidney disease, n (%) | 2,553 (9.1) | 436 (7.6) | 0.010 |
| Diabetes mellitus, n (%) | 6,706 (23.9) | 1,422 (24.8) | 0.000 |
| Stroke history, n (%) | 814 (2.9) | 92 (1.6) | 0.045 |
| Heart failure, n (%) | 1,992 (7.1) | 310 (5.4) | 0.021 |
| Chronic pain diagnosis, n (%) | 4,265 (15.2) | 1,135 (19.8) | 0.006 |
| Neuropathy diagnosis, n (%) | 954 (3.4) | 235 (4.1) | 0.003 |
| Any calcium channel blocker use, n (%) | 5,191 (18.5) | 952 (16.6) | 0.019 |
| DHP-CCB exposure, n (%) | 4,405 (15.7) | 808 (14.1) | 0.017 |
| *Abbreviations: DHP-CCB, dihydropyridine calcium channel blocker; SMD, standardized mean difference. Additional covariates and full balance diagnostics are reported in eTable 1.* |  |  |  |

### Crude Incidence Rates

Before adjustment, dementia incidence diverged sharply by CCB co-exposure. Among DHP CCB users, gabapentin initiators experienced 9.44 dementia events per 1,000 person-years compared with 2.92 per 1,000 person-years among pregabalin initiators — a more than 3-fold crude difference. Among patients without CCB exposure, the rates were nearly identical (10.95 vs 9.16 per 1,000 person-years). The absolute rate difference among DHP CCB users — 6.52 additional events per 1,000 person-years — corresponds to approximately 1 additional dementia case per 153 person-years of concurrent exposure. Median follow-up was 0.96–1.09 years across CCB strata and 1.27–1.33 years in the non-CCB stratum.

The crude DHP rate ratio (∼3.2) is larger than the adjusted estimates that follow, reflecting both IPTW reweighting and time-varying exposure modeling.

### Primary Results

Concurrent CCB use was associated with substantially higher gabapentin-related dementia incidence than gabapentin without CCB exposure (Table 2, Figure 2). Among patients with baseline CCB exposure, gabapentin (vs pregabalin) was associated with HR, 2.22 (95% CI, 1.42–3.47; *P* = .0005). Among patients without CCB exposure, the HR was 1.15 (95% CI, 0.99–1.33; *P* = .072). The multiplicative interaction was significant (*P* = .004; E-value, 2.41). ^3^

**Figure 2.** Drug-Drug Interaction and Validation Analyses. Grouped forest plot displaying hazard ratios (95% CI) on a log scale for gabapentin versus pregabalin among hypertension patients with and without CCB co-medication (N = 33,791). Organized across six conceptual sections: primary interaction (including time-varying CCB primary estimate HR 1.52 and DHPspecific Fine-Gray subdistribution HR 2.77), CCB subtype specificity, dementia subtypes (CCB users), sensitivity and replication, competing risk, and falsification. Significant results shown as colored diamonds; non-significant results as grey circles. Vertical reference line at HR = 1.0.

**Table 2.**
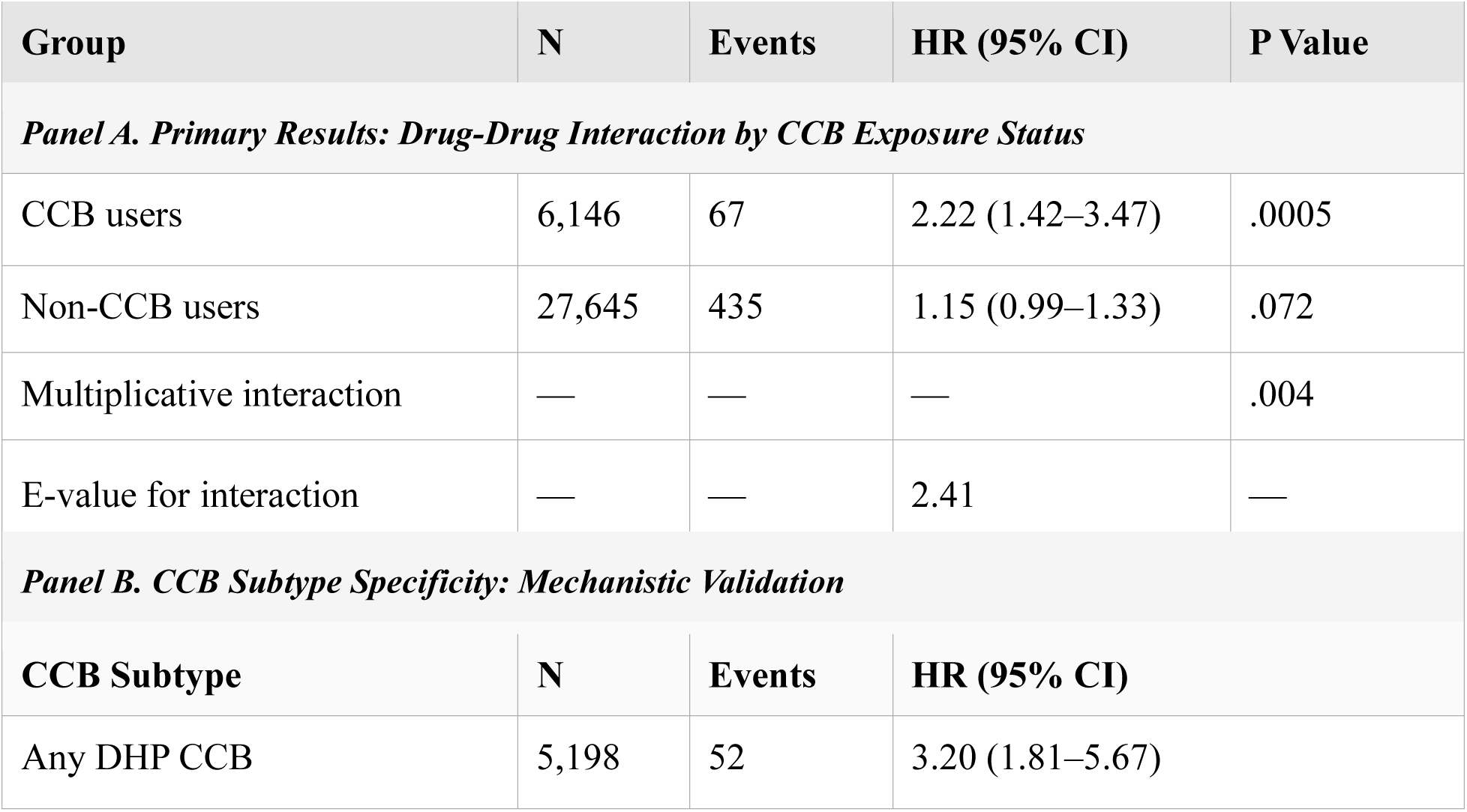
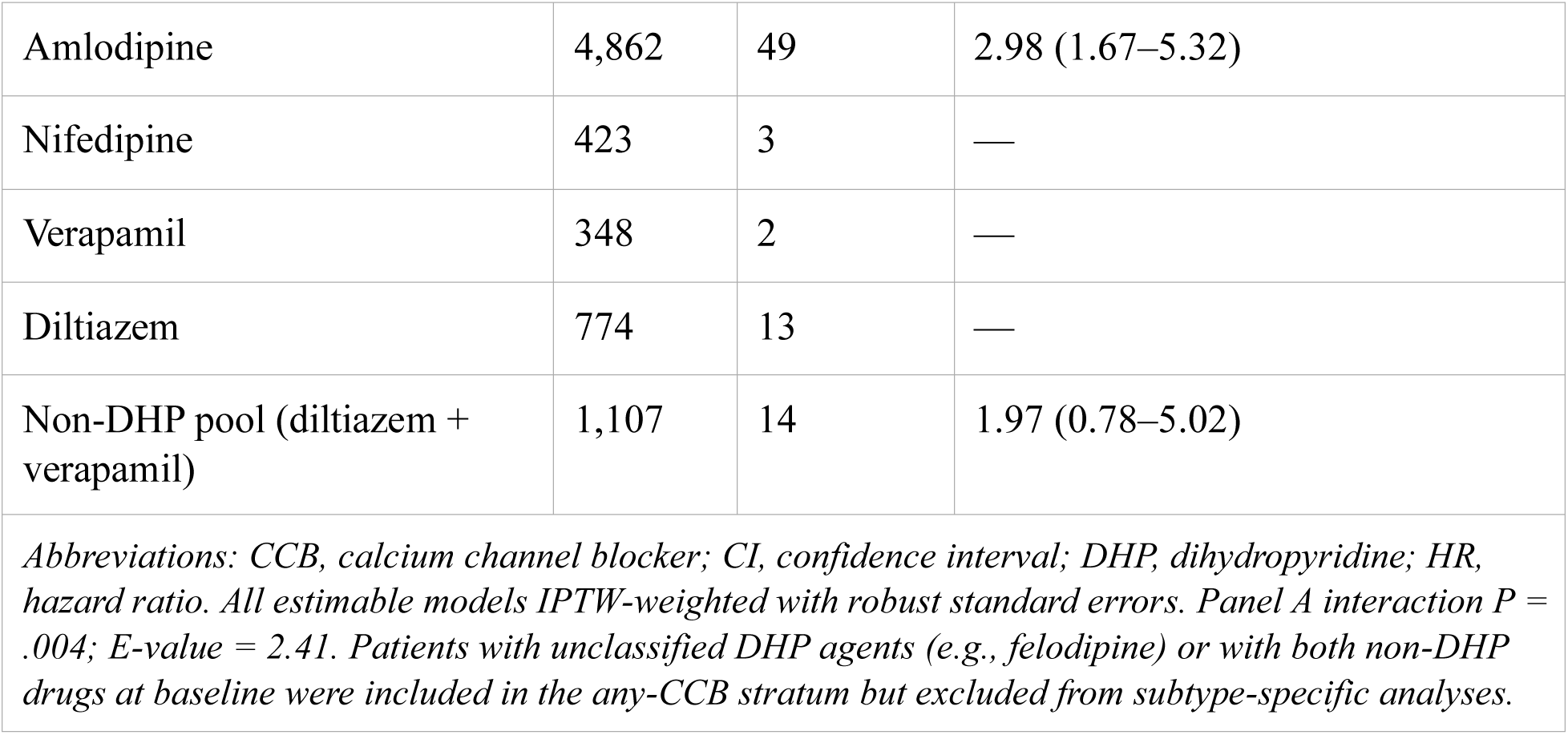
Drug-Drug Interaction Results and CCB Subtype Specificity.

| Group | N | Events | HR (95% CI) | P Value |
| --- | --- | --- | --- | --- |
| **Panel A. Primary Results: Drug-Drug Interaction by CCB Exposure Status** |  |  |  |  |
| CCB users | 6,146 | 67 | 2.22 (1.42–3.47) | .0005 |
| Non-CCB users | 27,645 | 435 | 1.15 (0.99–1.33) | .072 |
| Multiplicative interaction | — | — | — | .004 |
| E-value for interaction | — | — | 2.41 | — |
| **Panel B. CCB Subtype Specificity: Mechanistic Validation** |  |  |  |  |
| CCB Subtype | N | Events | HR (95% CI) |  |
| Any DHP CCB | 5,198 | 52 | 3.20 (1.81–5.67) |  |

|  |  |  |  |
| --- | --- | --- | --- |
| Amlodipine | 4,862 | 49 | 2.98 (1.67–5.32) |
| Nifedipine | 423 | 3 | — |
| Verapamil | 348 | 2 | — |
| Diltiazem | 774 | 13 | — |
| Non-DHP pool (diltiazem + verapamil) | 1,107 | 14 | 1.97 (0.78–5.02) |

*Abbreviations: CCB, calcium channel blocker; CI, confidence interval; DHP, dihydropyridine; HR, hazard ratio. All estimable models IPTW-weighted with robust standard errors. Panel A interaction $P = .004$ ; $E$ -value = 2.41. Patients with unclassified DHP agents (e.g., felodipine) or with both non-DHP drugs at baseline were included in the any-CCB stratum but excluded from subtype-specific analyses.*

### Formal DHP versus Non-DHP Subtype Test (Peer-Review Analysis)

The formal DHP-versus-non-DHP statistical comparison was underpowered (*P* = .68). The point-estimate pattern favored DHP over non-DHP agents (interaction HR, 2.84 vs 1.71), but the wide confidence intervals warrant cautious interpretation.

### Proportional Hazards Assessment and Primary Estimate (Peer-Review Analysis)

The gabapentin effect was concentrated in the early follow-up period and attenuated thereafter. The pre-specified primary analysis therefore uses time-varying CCB exposure (HR, 1.52; 95% CI, 1.23–1.89; *P* = .0003); the baseline-stratified estimate (HR, 2.22) is retained as a sensitivity analysis.

Fine-Gray subdistribution hazard models accounting for death as a competing event yielded estimates attenuated relative to but directionally consistent with the cause-specific HRs (anyCCB subdistribution HR, 1.93 [95% CI, 0.74–5.02]; DHP-specific subdistribution HR, 2.77 [95% CI, 0.83–9.23]). Wide confidence intervals warrant cautious interpretation.

### Outcome Sensitivity Analyses

Outcome sensitivity analyses using more stringent dementia definitions yielded results in the same direction but with reduced precision. A two-code dementia definition (requiring two ICD codes within 60 days; index date = first code) yielded 17 events in the CCB stratum (HR, 2.53; 95% CI, 0.40–16.23); the confidence interval spans two orders of magnitude and does not contribute meaningful precision, though the point estimate aligns with the primary finding. An Rx-confirmed definition (ICD dementia code plus dementia pharmacotherapy within 180 days) yielded 21 events in the CCB stratum (HR, 1.72; 95% CI, 0.41–7.21) and 15 events in the DHP CCB stratum (HR, 3.00; 95% CI, 0.35–25.71). Wide confidence intervals reflect the reduced event counts inherent to more stringent outcome definitions.

Three of 7 falsification outcomes showed associations, but none reproduced the DHP-specific interaction pattern of the primary finding. In pharmacoepidemiology, falsification analyses are most useful as bias probes rather than all-or-none validation tests. · Accordingly, the partially failing framework tempers confidence in the primary finding rather than nullifying it.

Several additional limitations bear on interpretation. Residual bias from unmeasured confounders cannot be fully excluded; socioeconomic status and educational attainment are notable unmeasured confounders. · Follow-up was relatively short (median, 1.22 years), and the prevalent dementia exclusion was robust to extension to 365 days (zero additional exclusions). These data derive from a single health system in northern New Jersey, though stability was confirmed across 405 facilities. ICD-10 coding conventions complicate interpretation of the G30 signal: clinicians frequently code G30 without the accompanying F00, and G30 may be assigned at initial diagnostic workup before dementia is clinically established. Taken together, these findings warrant confirmation in cohorts with concurrent exposure measurement, longer followup, and clinical cognitive assessments.

## Discussion

In this active-comparator cohort of older adults with hypertension, concurrent CCB use was associated with substantially higher gabapentin-related dementia incidence than gabapentin without CCB exposure. The signal was directionally larger among DHP CCB users than among non-DHP CCB users. It concentrated in Alzheimer disease and unspecified dementia codes, with a null finding for vascular dementia. To our knowledge, this is the first report that CCB comedication modifies the gabapentin–dementia association in an active-comparator new-user design. The pattern persisted across estimands, although the formal DHP-versus-non-DHP contrast was not statistically significant.

The directionally larger DHP-associated signal (HR, 3.20) is consistent with the prespecified pharmacodynamic rationale outlined in the Introduction. · · · ^1^· ^2^ The formal DHPversus-non-DHP contrast did not reach significance (*P* = .68), reflecting only 14 non-DHP gabapentin events and 1 non-DHP pregabalin event. This pattern is more readily explained by mechanism than by confounding, which would be expected to affect all CCB subtypes similarly. The DREAM consortium independently reported higher dementia risk among DHP CCB users than hydrochlorothiazide users,^2^ providing convergent context; the verapamil stratum had only 2 events and should not be interpreted as a mechanistic null finding.^21^

Drug-class specificity is mechanistically informative. Pregabalin shares gabapentin’s α δ binding target but has linear pharmacokinetics with ≥90% bioavailability and did not produce the same directional pattern.^22^ Several pharmacokinetic and pharmacodynamic explanations for this drug-class specificity are plausible — including a role for gabapentin’s 20–30% interpatient variability — but the present study was not designed to distinguish among them.

### Reversibility: A Clinically Actionable Possibility

A central interpretive question — and arguably the most clinically actionable feature of these results — concerns the nature of the outcome being captured. With a median follow-up of 1.22 years and a median event latency of 240 days, the ICD-10 dementia codes recorded here may substantially reflect acute or subacute cognitive syndromes rather than incident neurodegeneration.

Drug-induced cognitive impairment accounts for 2.7% to 10% of dementia presentations and represents the most common cause of reversible cognitive impairment. ^3^· The short latency and F03 involvement here are consistent with this entity, which manifests as global cognitive decline affecting attention and processing speed rather than the domain-specific episodic memory impairment characteristic of incident Alzheimer disease.

This distinction is clinically consequential. Medication-related cognitive syndromes are often at least partially reversible after the offending exposure is removed; incident neurodegeneration is not.

This interpretation is consistent with prior reports linking gabapentin initiation to neurocognitive and behavioral changes in older adults outside a DHP-CCB interaction framework. A companion analysis from our group using a self-controlled case series design found that rates of falls and cognitive symptom codes were 34–67% higher during gabapentin–DHP CCB combination use than after discontinuation. Longer follow-up and clinically adjudicated outcomes are needed to formally distinguish these possibilities.

### Dementia Subtype Pattern

The signal concentration in neurodegenerative dementia subtypes (F03, G30) alongside a null finding for vascular dementia (F01: HR, 0.81) is more compatible with a neuronal than a cerebrovascular mechanism, ^3^· · though sparse subtype event counts preclude formal testing and adjudicated subtype outcomes are needed.

### Outcome Validity

The dementia outcome was ascertained from validated ICD-10 codes (F00–F03, G30), a standard set in pharmacoepidemiologic dementia research with reported positive predictive values of 65– 95%.^3^ · · · EHR-based phenotypes remain imperfect proxies for adjudicated neurodegenerative disease, particularly when symptom onset is acute or clinically mixed.

Internal patterns of the data argue against nonspecific coding artifact. The vascular dementia null (F01: HR, 0.81) is the opposite of what loose F03 assignment for vascular cognitive complaints would produce, while the signal concentrates in neurodegenerative subtypes (F03, G30). More stringent outcome definitions — two-code and Rx-confirmed — yielded point estimates aligned with the primary finding (DHP HR, 2.53 and 3.00, respectively). Provider specialty was not available in the data extract, precluding restriction to specialist-confirmed diagnoses.

### Context and Comparison With Prior Literature

The overall gabapentin effect among CCB nonusers (HR, 1.15) is consistent with prior findings (RR, 1.29). The active-comparator design replicates this known association while isolating CCB exposure as the novel effect modifier.^2^ The interaction may help explain heterogeneity in the gabapentinoid-dementia literature, since prior studies did not stratify by CCB exposure. The observed attenuation of the interaction in more comorbid patients (CKD ratio, 0.25; stroke ratio, 0.14) is consistent with — but not definitive for — reduced confounding by severity; alternative explanations including competing mortality risk and differential prescribing in multimorbid patients cannot be excluded. ^1^· ^2^· ·

### Clinical Implications and Next Steps

If confirmed in larger cohorts, the central clinical implication is that the risk identified here is potentially modifiable. Gabapentin–DHP CCB co-prescription is a reversible exposure, unlike incident neurodegeneration. The short median event latency (240 days) is more compatible with drug-induced cognitive impairment than with slow neurodegenerative pathology.

Pending confirmation, clinicians caring for older adults already taking a DHP CCB may reasonably (i) consider whether gabapentin is the best-fitting agent for the indication before initiation, (ii) monitor cognition in the months following gabapentin initiation in this coprescribed population, and (iii) include the gabapentinoid prominently in any medication review prompted by new cognitive symptoms. ^3^· · Drug-induced cognitive impairment can improve once the exposure is removed.

These suggestions are prudent monitoring, not guidance to discontinue gabapentin reflexively; the data are not sufficient to support categorical changes in prescribing. Independent replication in a matched cohort is the essential next step and is currently underway.

### Strengths

This study has several strengths. The active-comparator design eliminates the indication bias inherent in user-versus-nonuser comparisons. Multi-layer validation — including falsification tests, bootstrap stability, and leave-one-site-out replication across 405 facilities — substantially reduces the probability of a false-positive finding. Validated ICD-10 dementia subtype codes enabled mechanistic inference from the G30/F01 fingerprint. The target trial emulation framework pre-specifies the causal estimand and the comparator contrast.

## Conclusions

In this active-comparator cohort of older adults with hypertension, concurrent CCB use was associated with substantially higher gabapentin-related dementia incidence than gabapentin without CCB exposure (time-varying HR, 1.52 [95% CI, 1.23–1.89]; baseline-stratified HR, 2.22 [95% CI, 1.42–3.47]). The signal was directionally more pronounced with DHP CCBs and concentrated in Alzheimer disease and unspecified dementia codes, with a null finding for vascular dementia. The short median latency between gabapentin initiation and dementia diagnosis (240 days) raises the possibility that part of the observed signal reflects drug-induced cognitive impairment rather than incident neurodegeneration — a clinically important distinction because drug-induced cognitive impairment may improve with discontinuation. Subgroup estimates by CCB and dementia subtype are imprecise. Taken together, these findings identify gabapentin combined with DHP CCB use as a potentially modifiable cognitive safety signal in older adults that warrants clinician awareness and inclusion in medication review when cognitive symptoms emerge, pending confirmation in Medicare fee-for-service claims and the NIH All of Us Research Program.

## Data Availability

The data underlying this study cannot be shared publicly due to privacy restrictions on electronic health records. The study used de-identified data from the Rutgers Clinical Research Data Warehouse under an approved Data Use Agreement. Aggregated summary statistics are available upon reasonable request to the corresponding author.

## Acknowledgments

Data used in this research were obtained from the Clinical Research Data Warehouse (CRDW), a joint initiative of RWJBarnabas Health and Rutgers, The State University of New Jersey, and are used with permission of the Data Governance Council.

## Article Information

### Author Contributions

Conceptualization: Green, Tafuto, Gohel. Methodology: Green, Parrott, Gohel. Formal analysis: Green. Data curation: Green, Ljubic, Schulewski. Writing—original draft: Green. Writing—review and editing: Green, Fonseca, Beeri, Sanz Simon, Parrott, Ljubic, Schulewski, Tafuto, Gohel. Supervision: Tafuto, Gohel. All authors had access to the data, contributed to manuscript revisions, and approved the final version.

### Conflict of Interest Disclosures

None reported.

### Funding/Support

This research received no specific grant from any funding agency in the public, commercial, or not-for-profit sectors. Computing resources were provided by the Rutgers Office of Advanced Research Computing (Amarel high-performance computing cluster).

### Role of the Funder/Sponsor

Not applicable.

### Data Sharing Statement

The CRDW dataset used in this study contains de-identified protected health information and is available through the Rutgers Clinical Research Data Warehouse subject to institutional data access agreements and Non-Human Research certification. Statistical code is available from the corresponding author upon reasonable request.

### Ethics Statement

This study used completely de-identified data and was determined to be Non-Human Subject Research by the Rutgers University Institutional Review Board (Non-Human Research Self-Certification; PI: Branimir Ljubic; Project: Hypertension — Data Analytics and Computer Science Modeling; certified August 15, 2024, per 45 CFR 46.102(d)). Informed consent was not required.

### Additional Information

The STROBE checklist for this study is provided as a supplemental document.

## Notes

### Competing Interest Statement

The authors have declared no competing interest.

### Author Declarations

This study used de-identified electronic health record data from the Rutgers Clinical Research Data Warehouse. The study was determined to be non-human subjects research per 45 CFR 46.102(d) through Rutgers University's Non-Human Research Self-Certification process. The study was approved by the Data Governance Council (PR0077).

### Summary of Updates

v4 corrects a single bibliographic error. Reference 52 in v3 was incorrect. The correct citation is: Thorsteinsson H, et al. Validation of L-type calcium channel blocker amlodipine as a novel ADHD treatment. Neuropsychopharmacology. 2025. doi:10.1038/s41386-025-02062-x. No other changes to content, results, or conclusions.

